# Residential Methamphetamine Contamination in Boulder Colorado: Incidence and Implications

**DOI:** 10.1101/2024.07.28.24311138

**Authors:** James E. Dennison, Norris Minick

**Affiliations:** Century Environmental Hygiene, Fort Collins, CO, USA; Agents for Home Buyers, Boulder, CO, USA (ret.)

**Keywords:** drug, exposure, methlab, thirdhand

## Abstract

The manufacture and use of methamphetamine (meth) is a significant problem, in part because it can lead to the contamination of properties where it occurs. Meth contamination can lead to health issues for occupants as well as very high remediation costs for property owners. But even in a state like Colorado, where meth testing and remediation are highly regulated, the number of residences or other types of property that are contaminated in excess of health standards is unknown. Generally, testing for meth contamination occurs only after a property is identified as a likely site for meth use or manufacture, whether by law enforcement, the property owner, or a potential buyer. For this paper, a unique random data set based on a real estate broker’s meth screening program was used to determine the incidence of contaminated residences in general. Brokerage clients put 303 residential properties under contract between 2013 and 2022, of which 288 (95%) were screened for meth contamination. Meth was detected in 45 of the 288 screening tests (16%), many at trace levels, while 84% contained no detectable meth. Comprehensive testing was subsequently performed on 35 of these 45 properties and ten of the original 288 (3.47%) screened properties contained meth contamination above state health standards. While the sample size of this analysis was modest, it provides a first real estimate of the incidence of meth-contaminated residential property and an indication of the environmental health significance of this issue.

## INTRODUCTION

Meth use has been a growing problem in the U.S. since the 1980’s (Gonzales et al. 2010; SAMSHA 2017, 2018, 2019, 2020, 2023). Buildings in which meth is manufactured (“cooked”) or used can readily become contaminated with the drug (Martyny et al. 2008; VanDyke M 2009) which has raised concerns in the public/environmental health sector regarding potential exposure and health consequences to non-users (McGuinness and Pollack 2008; Castaneto et al. 2013). Meth is frequently detected in hair, oral fluids, and urine in environmentally exposed children (Farst et al. 2011; Bassindale 2012; Castaneto et al. 2013). In a study of people living in contaminated residences with no reported on-going meth cooking or use, meth was detected in 56% of the occupants’ hair (Wright et al. 2020). Public health agencies in some locations have promulgated meth health standards or guidelines for the concentration of meth deemed acceptable for occupants third-hand exposures (ACC 2011; NewZealand 2017; MLCC 2024). Since universal testing of residences for meth is not required and is not routinely performed, the magnitude of the problem has been essentially unknown. In order to understand the number of people exposed to concentrations above health standards, the number of residences or other property that contain meth contamination is needed.

Meth remediation of buildings is quite costly, depending on applicable regulatory requirements and standards. In Colorado, state regulations require testing to be performed by licensed industrial hygienists and remediation to be performed by licensed meth contractors. Procedures and standards are stringent, and thus the cost of a testing/remediation project for a meth-contaminated house is generally in the tens of thousands of U.S. dollars, with total cost to owners often exceeding $50,000 for a project. This is often a significant percentage of asset value, and is therefore a concern for owners and investors, as well as landlords, property managers, or others.

Testing is performed for several different reasons, including but not limited to:

1. In Colorado, if law enforcement finds evidence of meth cooking or use, and informs the owner, the owner is required to test (C.R.S. 2024).
2. If the owner otherwise has knowledge that meth was cooked or used, testing is also required (C.R.S. 2024). In either case, testing is often, although not always, performed. Events 1 and 2 trigger a compliance test that costs $1,000 or more for a typical house.
3. At times, an owner, property manager, or other person has concern but no specific evidence that meth activity occurred in a property and conducts a test to know one way or the other.
4. Some buyers of real estate, including single family homes, condominiums, or apartment-style buildings conduct a due diligence test prior to purchase. In general, property buyers are more likely to commission a test when there are indications of risk. Indicators of contamination risk include rumors of drug use, poor condition, gang graffiti, etc. As Events 3 and 4 are voluntary, less expensive test protocols may be used.

To be declared in compliance with the health standard after a trigger event, a property in Colorado must be tested in a Preliminary Assessment (PA) with a defined sampling protocol. This test typically costs $1,000 - $3,000 per single home and is rarely performed during purchase/sale real estate inspections due to the cost. As described below, the initial tests that were done on properties in the dataset used here were screening tests, usually Targeted Meth Screenings (TMS). These include a more limited number of samples than a PA, and deliberately provide a positively biased result that triggered further PA testing if meth was detected.

No previous random surveys for the incidence of contaminated property were identified in the published literature. A previous assessment of meth incidence was developed for Australia (Parker and Howell 2021). The assessment relied on statistical meth use data within the population as opposed to random in-house testing data. It assumed that one meth user would contaminate one property to a detectable level but not necessarily above the Australian health standard (which is coincidentally or not coincidentally the same as the Colorado standard). No basis for the one user-one contaminated property was available. Interestingly, the assessment concluded that 1.5% of Australian homes may be contaminated to some degree. The present study seeks to extend this approach by determining the numbers of residences that are expected to exceed regulatory compliance limits based on measured meth concentrations in randomly selected homes.

In 2010, a real estate brokerage in Boulder, Colorado (Agents for Home Buyers (A4HB)) learned from a newspaper article that the home a client was set to close on the following week may have been contaminated by meth smoking. The broker commissioned a meth test which confirmed the property was contaminated, and the buyer was spared from buying a contaminated property. Unfortunately, numerous cases of buyers unwittingly purchasing contaminated properties have been reported in the news (Brosher 2015; Trapasso 2019; CBS 2020).

Thereafter, A4HB began encouraging buyers to screen houses for meth. From 2011-2012, A4HB tested 28 homes (50-75% of the homes put under contract) and 10% of these were found to have detectable meth in them. Starting in 2013, virtually all A4HB clients began to do meth screenings as part of the inspection. The TMS on its own would not define whether a property was in compliance, but the majority of the properties that had detectable meth in the TMS were followed with a PA. The approach in this assessment used the combination of data from the two test types to determine the incidence of legally contaminated houses.

## MATERIALS AND METHODS

“Contamination” can be viewed in two ways: (1) any detectable finding of meth or (2) finding meth at concentrations exceeding the applicable health standard. In Colorado, the health standard for meth in most areas is 0.5 ug/100 cm^2^ of surface area. Other states that have health standards have set them between 0.05 and 1.5 ug/100 cm^2^ (MLCC 2024). For the purposes of this paper, however, “contaminated” means “contaminated in excess of Colorado’s health standard.”

From 2013 through 2022, A4HB clients put 303 homes under contract, and screened 288 of them (95%) for meth contamination. Most of the buyers who opted not to test were purchasing new homes, where risks seemed lower, and where the buyer had no contractual right to conduct inspections. All homes were located within about 35 miles of Boulder, with 23% in the city itself, 47% elsewhere in Boulder County, and 30% in the surrounding areas. They were all single-family homes, townhomes, or condominiums, and none were multiunit apartment buildings. This paper is based on the data between 2013 and 2022, where 95% of the houses were tested.

All of the meth sampling for the A4HB dataset was done by experienced industrial hygienists. A Boulder firm performed all but four of the tests, which were performed by one of the authors. Wipe samples for all testing were collected according to NIOSH Method 9111, which entails sampling 100 cm^2^ of surface area with cotton gauze wetted with isopropanol. To limit TMS costs, nearly all samples were composited, combining samples from four different locations. Sample locations were taken from areas that were likely to have the highest levels of contamination, including HVAC systems and exhaust fans in bathrooms, kitchens and other locations. Samples were analyzed by AIHA accredited commercial laboratories by NIOSH Method 9111. Four of the 288 screenings were done in a more robust manner, but this does not affect the outcome of the incidence calculations.

Of the 288 properties screened, 35 were subsequently tested using the full PA protocol by a variety of certified consultants. Data were extracted from the PA reports, which are publicly available. According to the meth regulations, a property is non-compliant, or contaminated, if any part of the property exceeds the health standard during a PA. A unique provision in the Colorado regulations as compared to most state meth regulations is that “limited exposure areas,” namely attics and crawlspaces, are subject to a higher standard of 4 ug/100 cm^2^. However, none of the properties failed merely due to concentrations in limited exposure areas; all non-compliant properties had at least one normal exposure area that exceeded the 0.5 ug/100 cm^2^ Colorado health standard. Also, two of the 35 follow-up tests were Screening Level Assessments (a specific Colorado protocol), which were permitted for these properties. The use of the Screening Level Assessment had no bearing on the outcome, as both properties were in compliance after this follow-up test.

The PA has a robust sampling protocol, consisting of a four-part composite in each room, including attics, crawlspaces, and detached structures, and four discrete samples from HVAC systems and some other items. The same sampling techniques were used in the PAs as in TMSs. All properties that had a result for any room or component that exceeded the health standard were deemed non-compliant.

## RESULTS

Of the 288 houses undergoing a meth screening, 45 (16%) were reported to have detectable meth present. Buyers terminated contracts on 7 of these 10 properties, so it is possible that these properties were taken off the market. Three properties were sold without further testing, with TMS results ranging from 0.013 to 0.33 ug meth/100 cm^2^. Of the 10 properties that did not undergo a PA, two had TMS results over 0.5 ug/100 cm^2^, and were at significant risk of failing in a PA, but are not counted as failures in calculations.

In the A4HB survey, 35 housing units were tested with a PA or Screening Level Assessment after the TMS reported detectable meth. Data for all positive TMS sites that underwent a subsequent PA are shown in Table 1. Ultimately, 10 of the original 288 housing units (3.47%) were found to be contaminated above Colorado’s compliance limits. According to the PA data, 16 (6%) of housing units in the area contain traces of meth less than the compliance limit of 0.5 ug/100 cm^2^. The TMS data suggest a somewhat higher incidence (13%) of trace (>0 and <0.5 ug/100 cm^2^) meth contamination levels due to the deliberate bias in the TMS protocol. Of the 35 houses that underwent a PA, 9 contained no detectable meth (3%).

**Table 1:** Results for all Positive TMS sites with follow-up testing.

| ID | High TMS | High PA | ID | High TMS | High PA |
| --- | --- | --- | --- | --- | --- |
| 234 | 0.018 | 0.015 | 261 | 2.2 | 0.91 |
| 32 | 0.028 | 0.018 | 288 | 1.3 | 1.3 |
| 235 | 0.048 | 0.02 | 16 | 7.8 | 2.84 |
| 257 | 0.018 | 0.024 | 26 | 0.13 | 10.8 |
| 22 | 0.05 | 0.03 | 260 | 0.23 | 18.3 |
| 8 | 0.03 | 0.05 | 11 | 5.75 | 23 |
| 3 | 0.08 | 0.05 | 21 | 60.5 | 182 |
| 10 | 0.03 | 0.06 | 28 | 432 | 4635 |
| 17 | 0.03 | 0.06 | 19 | 0.02 | BRL |
| 30 | 0.05 | 0.06 | 24 | 0.02 | BRL |
| 12 | 0.3 | 0.1 | 2 | 0.03 | BRL |
| 271 | 0.095 | 0.14 | 29 | 0.05 | BRL |
| 13 | 0.16 | 0.18 | 33 | 0.05 | BRL |
| 232 | 0.018 | 0.21 | 5 | 0.06 | BRL |
| 31 | 0.05 | 0.23 | 15 | 0.06 | BRL |
| 7 | 0.46 | 0.28 | 249 | 0.13 | BRL |
| 27 | 1.11 | 0.522 | 14 | 0.19 | BRL |
| 23 | 0.24 | 0.87 |  |  |  |

BRL: Below Reporting Limit (no meth detected)

It is of interest whether the TMS accurately predicts a PA result. Figure 1 shows the regression of the two test types with the two most extreme values omitted. A poor correlation is found between the two tests (R^2^ = .18). This is due to the high variability in the sampling method. It also shows a positive bias in the TMS, which is expected because the screening is designed to err on the positive side. However, inspection of the data reveals a low rate of false negatives in the TMS protocol.

**Figure 1:**
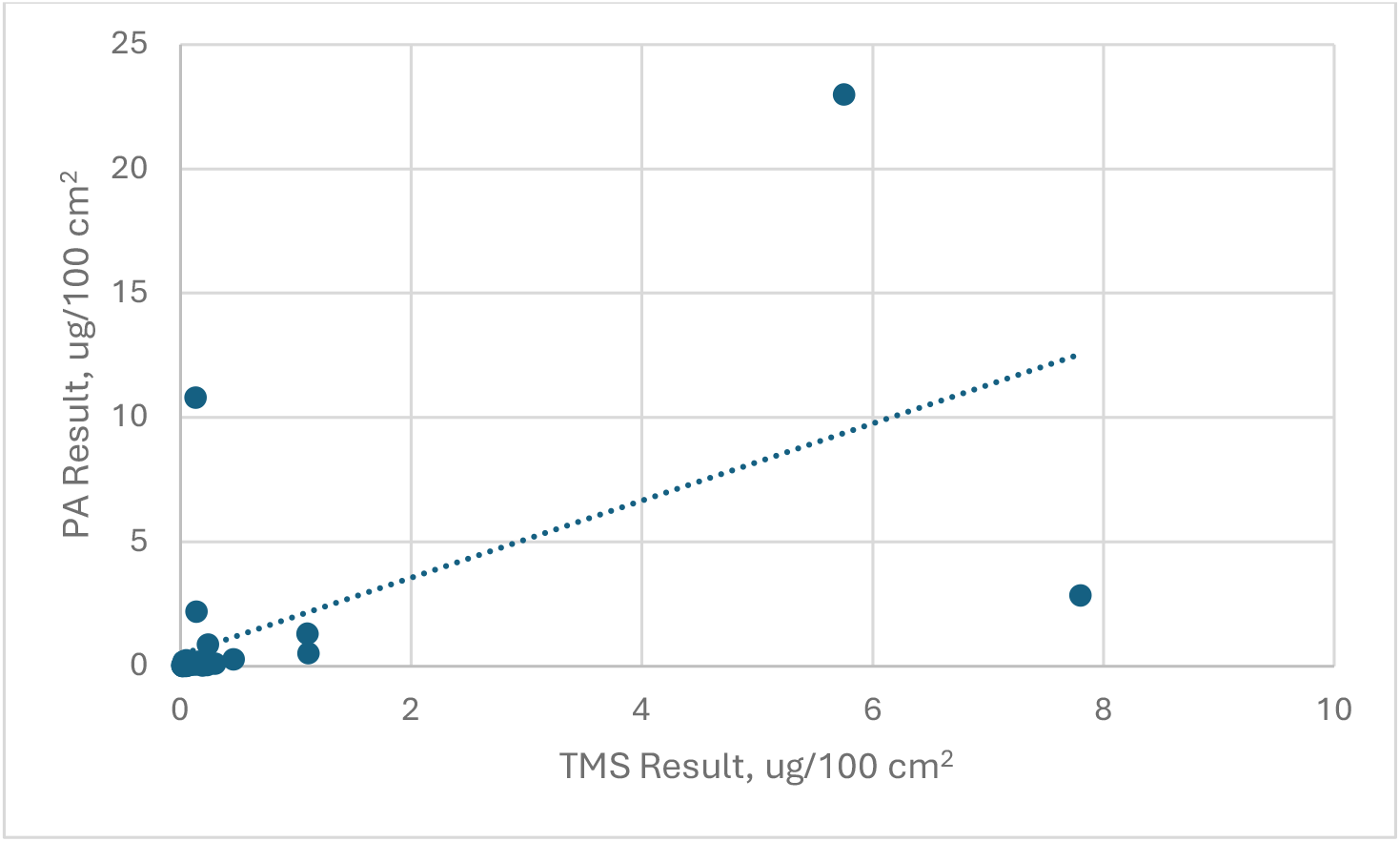
Regression of TMS and PA Results

## DISCUSSION

This analysis of the only identified random survey of meth incidence data suggests that a substantial number of housing units in the Boulder area are contaminated with meth. To put the A4HB data-based estimate of incidence into perspective, some non-random data may be worth noting. Colorado’s Neighborhood Stabilization state program operated in areas with high foreclosure rates, purchasing, renovating and selling 167 single family homes and 139 multifamily rental units. As reported in 2013, 7% of the single-family properties and 11% of the multifamily units in this program were contaminated (CNSP 2013).

In a dataset of one of the author’s separate work, a survey of 47 properties tested in 2008 provided the following results. Of the units tested, 72% were found to be contaminated. Of 24 houses that were in foreclosure status at that time, 75% were contaminated. Foreclosures would not be considered a random selection, but also not strongly biased, as buyers usually knew little about the property other than appearance. Rental properties in this dataset were contaminated 96% of the time.

During 2022-2023, one author supervised 37 TMSs not included in the A4HB survey. Forty-three percent of these had no detectable meth, 11% were identified with meth concentrations <0.5 ug/100 cm^2^, and 46% were identified with meth concentrations >0.5 ug/100 cm^2^. Of these 37 properties, 16 were subsequently tested with the PA protocol, and 14 failed in the PA (38% of the originally tested units). These three datasets are biased but show the potential that the actual percentage of contaminated units may be higher than in the A4HB dataset.

The A4HB data suggested that estimates of meth contamination incidence in housing based on biased “real-world” data sets like those discussed above grossly exaggerate actual contamination levels. This isn’t surprising, since PAs are generally performed only if a property has a high likelihood of contamination. The data also suggest that the Australian estimate of a 1.5% contamination rate may be low. This is really no surprise either, since many meth users will live in several homes while using meth, will smoke in properties they visit (e.g. friends and family), and even smoke in properties in which they work (e.g., house sitting or home remodeling).

### Uncertainty and Bias Analysis

PAs in the context of real estate sales were normally conducted only if a TMS detected meth. Some uncertainty exists in the ability of the TMS to avoid false negatives, i.e., meth was not detected in the TMS and if a PA were performed, would have had a result above 0.5 ug/100 cm^2^. A separate assessment of this error rate is underway, and indications are that the TMS had a very low rate of false negatives.

Errors in PAs could also occur (false negatives or positives.) Due to the high variability in surface residue concentrations, even composite samples do not always estimate the average concentration accurately. It is worth noting, though, that for all TMS with results >0.5 ug/100 cm^2^, none of the PAs reported the property to be in compliance.

Selection biases may exist. No expectation of bias in buyers’ decisions to be represented by A4HB exists. The small number of non-participants could reflect some concern that a house might test positive, but most were over newly built houses with a low (but not non-zero) probability of contamination. The A4HB survey augured against hidden bias by offering a discounted test to all buyer clients and strongly encouraging buyers to perform the test.

The error due to sample size cannot be estimated as there is only one random survey available. While a modest number of houses were screened for meth, only ten proved to be contaminated in this sample. Additional surveys will be needed to increase the accuracy of the estimate.

Neither the brokers, buyers, inspectors, or industrial hygienists who spent time in the homes tested for A4HB reported any signs of drug use, manufacture, or contamination. Also, very few of the properties were rental properties. Thus, the survey dataset contained a small negative bias in this regard. In addition, the 10 properties that were positive in the TMS and were not tested in a PA could reasonably have reduced the incidence estimate by 20-30%. These factors suggest that the real incidence rate could be higher than estimated.

The finding of 3.5% contaminated residences in the Boulder area may not be representative of the Boulder area nor of Colorado or the U.S. at large for other reasons, including:

1. The A4HB buyer properties did not include any housing units in apartment buildings, which may have a significantly higher incidence of meth contamination. The probability of meth use in a building is directly related to the number of residents over time and is also associated with occupant age. Since turnover in apartments is greater than in owned housing, and the occupants tend to be younger, apartments would be expected to experience more meth use.
2. Meth use has a higher association with lower socio-economic status (Tejiram et al. 2022). Boulder has higher median household income than the state and national average (U.S.CensusBureau 2024a). The average cost of housing units sold in the A4HB survey ($540K) was higher than the state average ($466K) and the national average ($282K) [9] but lower than the average price of homes sold in Boulder (Realtor.com 2024). Thus, the incidence of contaminated homes throughout the Boulder area may be lower than found here, but the incidence in Colorado and the U.S. at large might be higher based purely on socioeconomics.
3. The presence of a major university may also drive Boulder incidence rates down. Meth use is decreased among college students *vis a vis* same aged non-college adults (Johnston 2006).

Overall, it appears most likely that this survey has a negative bias compared to other regions.

It is difficult to obtain an unbiased measurement of the incidence of meth-contaminated houses. If a survey is contemplated for pure research purposes, difficulties in obtaining random participation would likely include concerns on the part of users over reporting of illegal activity, and the question of how contaminated properties would be mitigated after discovery. A buyer-initiated survey is a logical alternative to a government initiated one. Hopefully, these obstacles can be overcome so that broader-based surveys are done in the future.

### Does it make sense that many residential units are contaminated?

The process and speed at which residential buildings become contaminated are not well understood at this time. A single meth “cook” may contaminate a building to levels above health standards (VanDyke M 2009). Simulated meth smoking left surface residues of 0.2 – 0.9 ug/100 cm^2^ (John W. Martyny 2008; Russell et al. 2022), suggesting that a few episodes of meth smoking might contaminate to health standards. Within buildings, contamination appears to persist for a long time (Bitter 2017) with highly elevated concentrations found even years after the activity ceased.

### Effect of Health Standards

Promulgated state health standards in the U.S. range between 0.05 and 1.5 ug/100 cm^2^, a thirty-fold range (MLCC 2024). At this time, USEPA has not promulgated a standard or provided a numeric guideline (USEPA 2021). For jurisdictions with higher health standards, a lower percentage of homes will exceed standards and vice versa. Based on the A4HB dataset, the incidence of contaminated properties is shown in Figure 2 for a range of health standards. For a health standard of 0.05 ug/100 cm^2^, the data forecast 8% of properties would exceed the standard. For the standard initially promulgated by California (1.5 ug/cm^2^), 2% of properties are estimated to exceed the standard.

**Figure 2:**
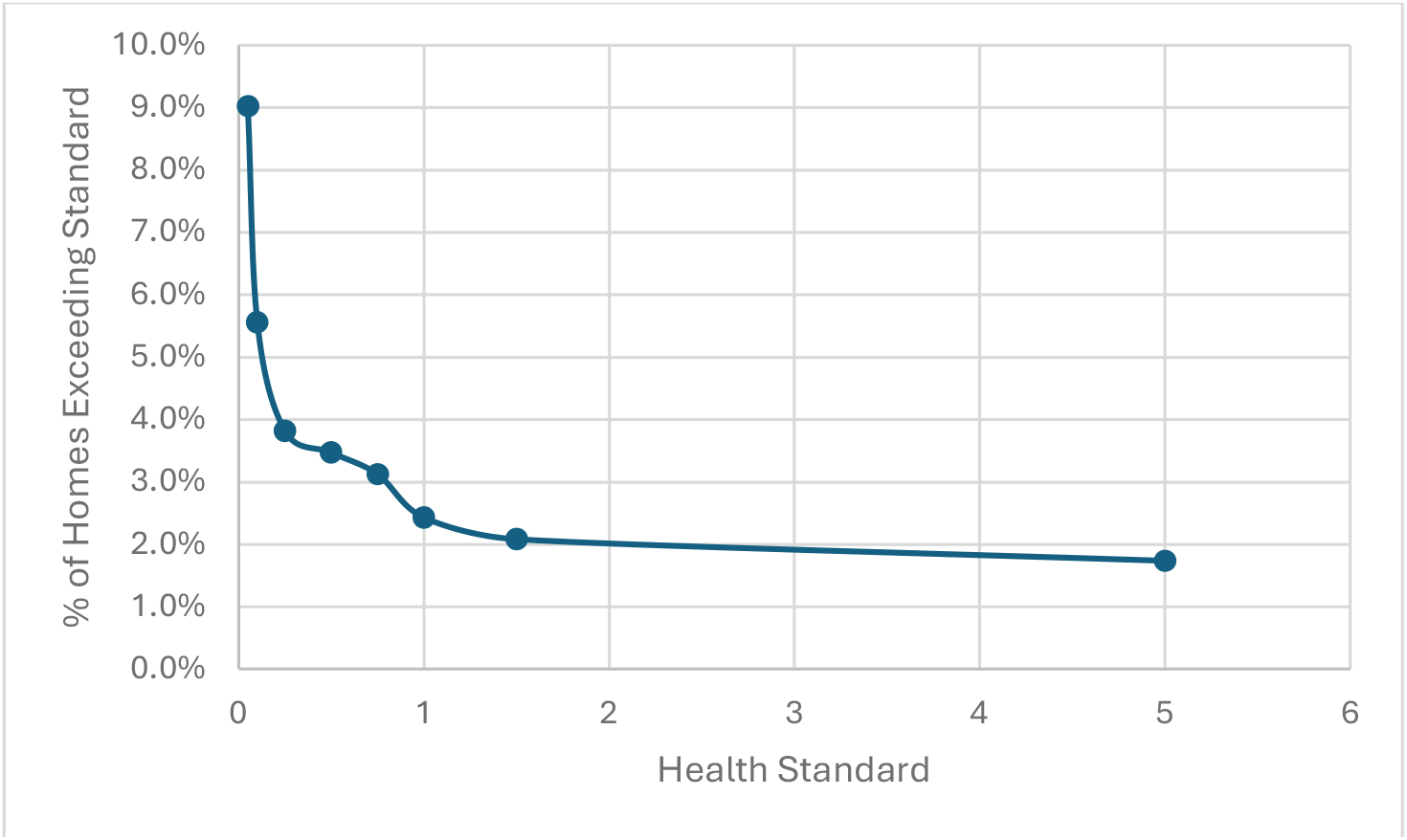
Percent of Properties Contaminated above Different Health Standards

### Public/Environmental Health Significance

The scope of this study did not include detailed assessment of the numbers of residences and people living in them that are expected to be contaminated. Further assessment of this is on-going. However, if 3.5% of Colorado’s population of 5.7 million people (U.S.CensusBureau 2024b) are in legally contaminated housing, over 200,000 people would be affected. At almost 100,000 single family homes sold each year in Colorado (Svaldi 2023), and less than 1% screened for meth, with a occupant density of 2.6 persons/house (WorldPopulationReview 2024), over 9,000 people are newly occupying houses with meth contamination in 3,500 houses they purchase annually. The magnitude of this issue clearly points towards a need to improve the estimates in this study with broader surveys.

## CONCLUSIONS

There is much to be learned regarding the incidence of meth-contaminated properties in the U.S. Broader random surveys should be performed that would allow accurate determinations of the numbers of contaminated properties throughout the U.S., and the rate of change in the numbers. It would also be important to determine the numbers of people residing in contaminated properties. The large disparity in state health standards suggests that the health risks are not well characterized and deserve re-evaluation. Finally, research into the rates and ways that meth contamination gets into properties, the fate of contamination within property, acute and chronic health effects, and more cost-effective remediation methods is needed.

## Data Availability

All data produced in the present study are available upon reasonable request to the authors

## Acknowledgements

The raw data for the A4HB database was used with permission from Mr. Minick and Lindsey Wolf Lunney.

